# High attack rates of SARS-CoV-2 infection through household-transmission: a prospective study

**DOI:** 10.1101/2020.11.02.20224485

**Authors:** Kanika Kuwelker, Fan Zhou, Bjørn Blomberg, Sarah Lartey, Karl Albert Brokstad, Mai Chi Trieu, Anders Madsen, Florian Krammer, Kristin GI Mohn, Camilla Tøndel, Dagrunn Waag Linchausen, Rebecca J. Cox, Nina Langeland, on behalf of the Bergen COVID-19 research group, Amit Bansal, Annette Corydon, Francisco Real, Geir Bredholt, Hauke Bartsch, Helene Heitmann Sandnes, Juha Vahokoski, Kjerstin Jacobsen, Marianne Eidsheim, Marianne Sævik, Nina Urke Ertesvåg, Synnøve Ygre Hauge, Therese Bredholt Onyango

## Abstract

**Background:** Household attack rates of SARS-CoV-2 ranging from 7% to 38% have been reported, using reverse transcription polymerase chain reaction (RT-PCR) of respiratory samples. Lower attack rates were described in children, but the importance of age in household transmission dynamics remains to be clarified.

**Methods:** During the first month of the outbreak, we enrolled 112 households (291 participants) in a prospective case-ascertained study, collecting demographic and clinical data from index cases and household members. Sera were collected 6-8 weeks after index case symptom onset, to measure SARS-CoV-2-specific antibodies.

**Findings:** T Local Ethics Committee (#118664). he overall household attack rate was 45% assessed by seroconversion, and 47% when also including RT-PCR positives. Serology identified a significantly higher number of infected household members than RT-PCR. Attack rates were equally high in children (43%) and young adults (46%), but highest among household members aged ≥60 years (72%). The attack rate was 16% in asymptomatic household members, and 42% in RT-PCR negative household members. Older adults generally had higher antibody titres than younger adults. The risk of household transmission was higher when the index case had fever or dyspnoea during acute illness but not associated with cough.

**Interpretation:** Serological assays provide more accurate estimates of household secondary attack rate than RT-PCR, especially among children who have a lower RT-PCR positivity rate. Children are equally susceptible to infection as adults, but elderly show higher attack rates. Negative RT-PCR or lack of symptoms are not sufficient to rule out infection in household members.

**Funding:** Helse Vest (F-11628), Trond Mohn Foundation (TMS2020TMT05).

## Introduction

Since first being identified in Wuhan, China in December 2019, the severe acute respiratory syndrome coronavirus 2 (SARS-CoV-2) has rapidly emerged into a global pandemic affecting over 180 countries. As of 5^th^ September 2020, there were more than 26 million confirmed cases of coronavirus disease 2019 (COVID-19), including over 870,000 deaths (1). In Norway, the first confirmed case of COVID-19 was identified on 26^th^ February 2020 (2). To combat further spread of the virus in the community, the government implemented comprehensive infection control measures on 12^th^ March 2020. However, quarantine of suspected cases and isolation of confirmed cases was practised from late February (3). As of 4^th^ September 2020, there have been 11,200 confirmed cases and 264 deaths in Norway (4).

Current testing for SARS-CoV-2 relies on amplification of the viral RNA genome from respiratory specimens, which can generally only be detected during acute infection. Whereas serological assays can determine exposure or infection over a longer time period, and are less dependent on the timing of sampling. Furthermore, with a high proportion of asymptomatic and mild infections (5), it is unlikely that data restricted to reverse transcription polymerase chain reaction (RT-PCR) provide the true infection rate of SARS-CoV-2.

The SARS-CoV-2 is a novel virus, and there are negligible levels of pre-existing antibodies in the population against the virus (6, 7). SARS-CoV-2-specific antibodies appear in the early convalescent phase approximately two weeks after infection and are maintained for at least four months (8, 9). Therefore, serological assays can provide valuable information on the real infection rate in a community. SARS-CoV-2 binds to the surface receptors of cells in the respiratory tract through the receptor-binding domain (RBD) on its spike protein, and neutralising antibodies prevent infections by blocking viral entry.

The secondary attack rate of SARS-CoV-2 from index cases to household contacts reflects the natural spread of infection in immunologically naive populations with limited preventive measures to control transmission. Respiratory tract infections have been documented to give varying secondary attack rates in families, particularly in influenza, where previous pandemics have reported rates from 4% to 20% or higher (10). With no immunity in the population, a higher household secondary attack rate would be expected with SARS-CoV-2.

Previous studies on the household transmission of SARS-CoV-2 have reported attack rates, ranging from 7·6% to 38%, based on RT-PCR of either single or repeated respiratory samples from household contacts of SARS-CoV-2 cases (11-20). However, sensitive serological assays are likely to give more accurate estimates of attack rates (21), regardless of whether household members are asymptomatic or RT-PCR negative.

Here, we estimated the secondary household attack rate of SARS-CoV-2 and identified the determinants of household transmission by measuring SARS-CoV-2-specific antibodies in household members of RT-PCR confirmed cases during the first month of the COVID-19 pandemic in Norway.

## Methods

### Study design, setting and participants

This prospective case-ascertained study was conducted in Bergen, Norway. Testing for SARS-CoV-2 by RT-PCR from throat swabs was centralized at Bergen Municipality Emergency Clinic for the city. All RT-PCR confirmed cases tested at the clinic during the start of the local outbreak (28th February–4th April 2020), and their household members were eligible for the study. Household members were defined as individuals who resided in the same household as a confirmed case, termed as index case hereafter. Index cases and their household members were contacted by telephone and asked to participate in the study. In households with >1 case, the member first diagnosed with COVID-19 was defined as the index case, and non-primary cases were defined as household members. Households where a case resided alone or no household members were willing to participate in the study, were excluded from the analysis. The study was approved by the Local Ethics Committee (#118664). All participants and/or their guardians provided written informed consent before inclusion in the study.

### Clinical information

Electronic case report forms (eCRF) were developed using REDCap® (Research Electronic Data Capture) (Vanderbilt University, Nashville, Tennessee). The eCRF for index cases contained demographics, COVID-19-like symptoms, recent travel history, recent close contact with confirmed COVID-19 cases, as well as household size and number of household members that had been ill with similar symptoms. Household members were contacted individually to register information on gender, age, RT-PCR test result (if available), and COVID-19-like symptoms.

### Serological assays

Serum samples from index cases and household members were collected 6-8 weeks after symptom onset in the index case. Sera were stored at −80°C and heat-inactivated for one hour at 56°C before use in serological assays.

### Enzyme-linked immunosorbent assay (ELISA)

A two-step ELISA was used for detecting SARS-CoV-2-specific antibodies, initially by screening with receptor-binding domain (RBD) and then confirming seropositivity by spike IgG (6) (see supplementary methods). Endpoint titres were calculated as the reciprocal of the serum dilution giving an optical density (OD) value=3 standard deviations above the mean of historical pre-pandemic serum samples (n=128) (supplementary figure 1). Individuals with no antibodies were assigned a titre of 50 for calculation purposes. Since the historical serum samples were defined as seronegative, and recruitment was initiated from the first case, we assume that all participants were seronegative at baseline and the term seroconversion is used for participants with seropositive spike-specific IgG.

### Neutralisation assays

The neutralisation assays were used to quantify SARS-CoV-2-specific functional antibodies (see supplementary methods). The assays were performed in a certified Biosafety Level 3 Laboratory using a Norwegian clinical SARS-CoV-2 virus at 2000 tissue culture infectious dose 50% (TCID_50_)/ml. In the microneutralisation (MN) assay, virus infectivity was measured by detecting the amount of nucleoprotein after 24 hours incubation in Vero cells. The MN titre (IC_50_) was determined as the reciprocal of the serum dilution giving 50% inhibition of virus infectivity. In the virus neutralisation (VN) assay, the cytopathic effect (CPE) in Vero cells was recorded after 4-5 days. VN titres were determined as the reciprocal of the highest serum dilution giving no CPE. Negative titres (<20) were assigned a value of 10 for calculation purpose.

### Statistical methods

Risk factors for seroconversion, including household member’s age, gender, RT-PCR-status and symptoms, the size of the household, and characteristics of the index case, were presented as proportions and assessed in univariable analysis by chi-square test for binary explanatory variables, and by logistic regression for factors with multiple levels, using the level with most observations as reference. A p-value of <0·05 was defined as the cut-off for statistical significance. Comparison of continuous variables, such as antibody titres, was performed by Mann-Whitney U test. Analyses were performed in R 4·0·2 (www.R-project.org). Graphs were drawn in Prism 7 (GraphPad Software, San Diego, CA)

### Role of the funding sources

The funding bodies had no role in study design, collection, analysis and interpretation of data, in writing the manuscript, and in the decision to submit this paper for publication.

## Results Participants

Between 28th February and 4th April 2020, 223 out of 3319 RT-PCR tested individuals were identified as SARS-CoV-2 positive in Bergen, and deemed eligible for the study. All positive cases were contacted, of which 194 cases were enrolled in the study. Among 258 eligible household members, 148 were enrolled in the study. In households where more than one case resided, the primary case was defined as the index case and 31 non-primary cases were redefined as household members. Fifty-one cases of single-person households were excluded from the analysis. The final sample used for analysis consisted of 112 index cases and 179 household members (figure 1 and 2B). Overall, there was an equal distribution of males and females in both index cases and household members, although household members were younger than the index cases (table 2). A large proportion (73%, 130/179) of household members reported having COVID-19 compatible symptoms (supplementary table 1).

**Figure 1.**
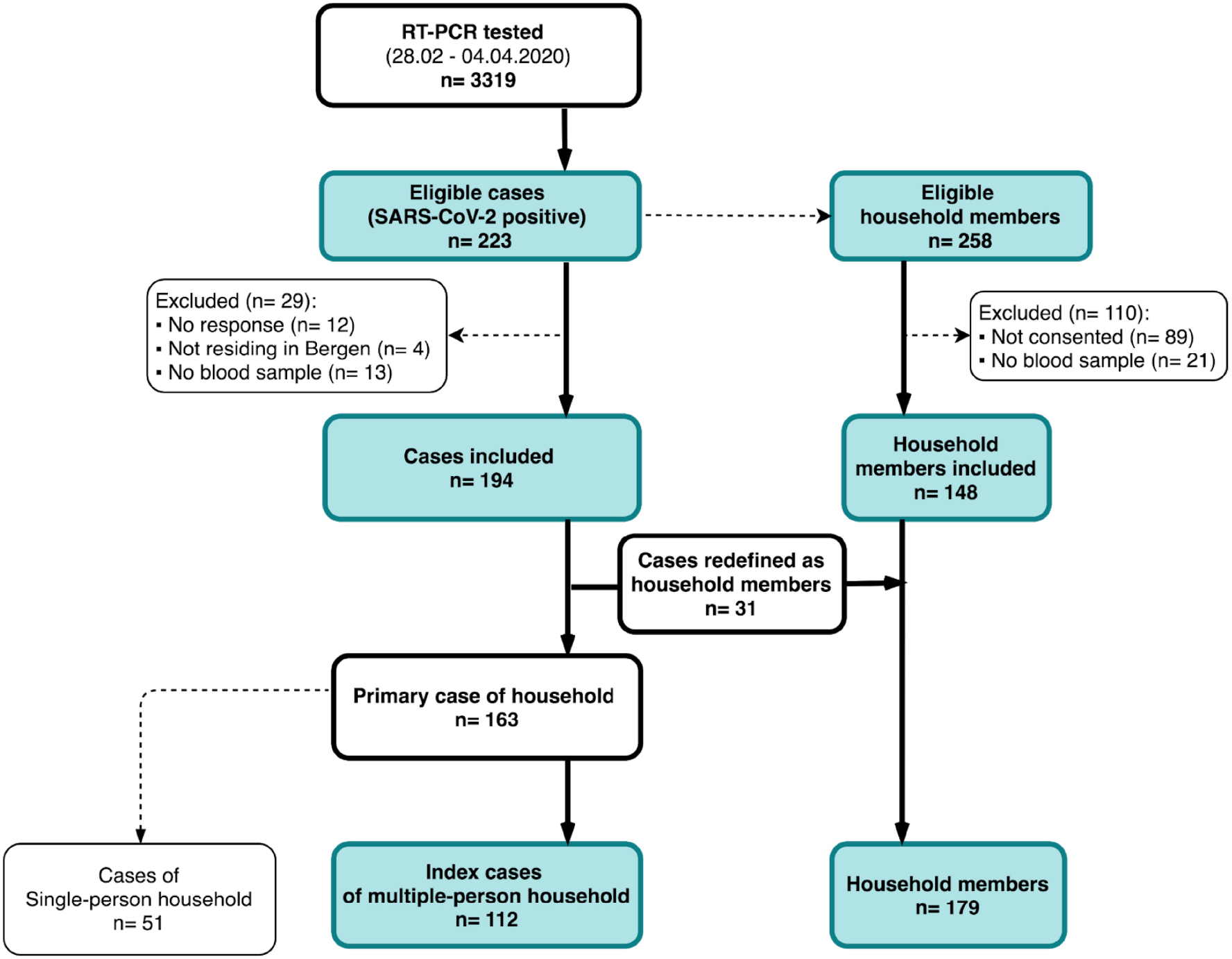
Recruitment procedure of study participants. The first case was diagnosed on 28^th^ February in Bergen, Norway’s second largest city. Between 28^th^ February and 4^th^ April 2020, 223 SARS-CoV-2 RT-PCR cases were identified among 3319 SARS-CoV-2 suspected cases that were tested at the Bergen Municipality Emergency Clinic, out of which 194 RT-CPR positive individuals were included in the study. There were 258 eligible household members in total, out of which 148 were included. In households with more than one RT-PCR positive case, 31 non-primary cases were redefined as household members. In total, we included 291 people comprising 112 index cases living with others and their 179 household members. Fifty-one cases were not included in the analyses as they lived alone or were defined as single person households because they did not have household members who wished to participate in the study.

**Figure 2.**
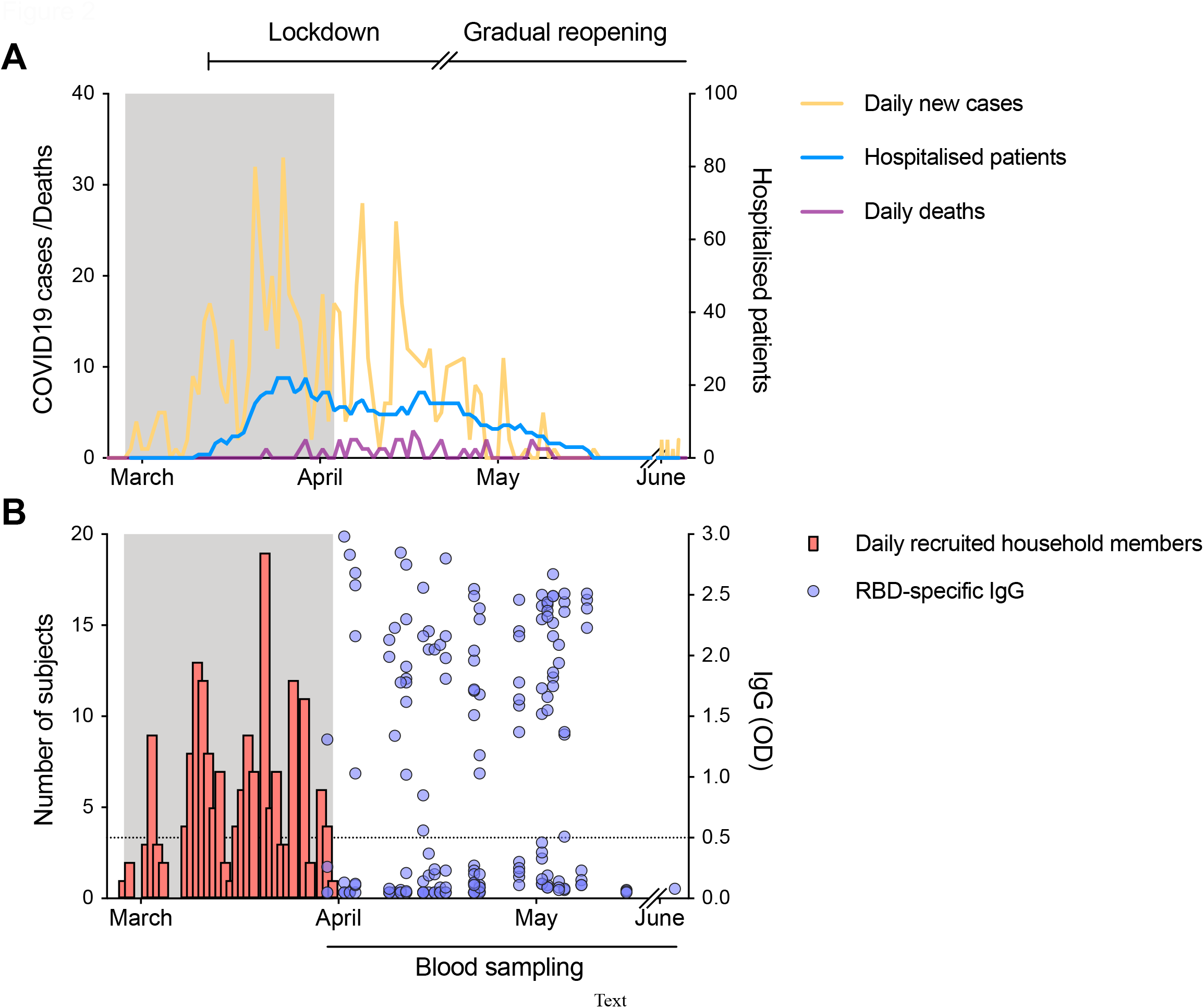
The course of the first wave of the pandemic in Bergen and period of recruitment of index cases and household members. The daily number of SARS-CoV-2-positive cases (shown in orange) from the centralised testing centre at Bergen Municipality Emergency Clinic covering a population of 284·000 people and the daily number of COVID-19 deaths (shown in purple) in Bergen, Norway (left Y axis). The number of hospitalised patients from SARS-CoV-2 infection in Bergen is shown in blue (right Y axis). Lockdown was initiated in Norway on 12th March, and a gradual reopening starting on 20th April 2020. **(B)** The number of household members recruited (shown in red) during the recruitment period (gray shaded area). Clinical information was collected from the index case and their household members at the time of recruitment. Blood samples were collected 6-8 weeks after the onset of symptoms in the index case (for one case serum was collected 3 weeks after inclusion, and for one case serum was collected 10 weeks after inclusion). Sera from all household members were tested against the receptor-binding domain (RBD) of spike protein in a screening ELISA. RBD-specific IgG are shown as the optical density (OD) at 1/100 dilution of sera (shown in blue, right Y axis). Each symbol represents one subject. The horizontal dotted line indicates OD 0.5 as the cut-off defined by a panel of 128 pre-pandemic sera. Duplicates were performed in ELISA.

### Attack rate defined by seroconversion

To calculate the attack rate, we measured SARS-CoV-2-specific IgG in household members using the spike protein ELISA to confirm seroconversion. The overall attack rate in households was 45%, with no significant gender difference (table 1). Attack rates varied between 25% and 72% among the different age cohorts. The elderly (≥60 years old) had a significantly higher attack rate (72%) than adults< 60years old (46%, p=0·045). Interestingly, the attack rate in children (43%) was similar to that of adults (46%). Similar trends were observed when using 10-year age cohorts with significantly higher attack rate in the oldest household members (supplementary table 2). Titres of spike-specific IgG among seropositive children <10 years old were significantly higher (p=0.03) than in adults (21-31 years old) (figure 3A). The presence of any COVID-19 symptoms among household contacts significantly increased the likelihood of infection (p<0.01), with seroconversion occurring in 56% of symptomatic and 16% of asymptomatic household members (table 1). At a household-level, household size was not convincingly associated with household transmission. Attack rates were highest in two-person households, but household size varied between age groups, with the majority of the oldest household members living in two-person households (table 1).

**Table 1:** Impact of household member characteristics on household secondary attack rate.

|  | n | Clinical symptoms<br>n (%) | P | n | Spike-specific IgG<br>n (%) | P | n | RT-PCR<br>n (%) | P |
| --- | --- | --- | --- | --- | --- | --- | --- | --- | --- |
| **Overall** | 179 | 130 (72·6) |  | 179 | 81 (45·3) |  | 70 | 32 (45·7) |  |
| **Sex** |  |  |  |  |  |  |  |  |  |
| Male | 80 | 57 (71·3) | 0·7 | 80 | 38 (47·5) | 0·6 | 34 | 15 (44·1) | 0·8 |
| Female | 99 | 73 (73·7) |  | 99 | 43 (43·4) |  | 36 | 17 (47·2) |  |
| **Age, years** |  |  |  |  |  |  |  |  |  |
| Children (0-12) | 28 | 20 (71·4) | 0·9 | 28 | 12 (42·9) | 0·8 | 9 | 1 (11·1) | 0·09 |
| Teenagers (13-19) | 24 | 15 (62·5) | 0·3 | 24 | 6 (25·0) | 0·07 | 5 | 2 (40·0) | 0·8 |
| Adults (20-59) | 109 | 79 (72·5) | ref | 109 | 50 (45·9) | ref | 47 | 21 (44·7) | Ref |
| Elderly ≥60 | 18 | 16 (88·9) | 0·2 | 18 | 13 (72·2) | **0·045** | 9 | 8 (88·9) | **0·04** |
| **Symptoms** |  |  |  |  |  |  |  |  |  |
| Yes | 130 | ·· | ·· | 130 | 73 (56·2) | **<0·001** | 66 | 32 (48·5) | 0·06 |
| No | 49 | ·· |  | 49 | 8 (16·3) |  | 4 | 0 (0·0) |  |
| **RT-PCR results\*** |  |  |  |  |  |  |  |  |  |
| Positive | 32 | 32 (100) | 0·06 | 32 | 28 (87·5) | **<0·001** |  | ·· | ·· |
| Negative | 38 | 34 (89·5) |  | 38 | 16 (42·1) |  |  | ·· |  |
| Not tested | 109 | 64 (58·7) |  | 109 | 37 (33·9) |  |  | ·· |  |
| **Household size** |  |  |  |  |  |  |  |  |  |
| 2 | 44 | 37 (84·1) | 0·06 | 44 | 25 (56·8) | 0·07 | 17 | 11 (64·7) | 0·09 |
| 3 | 36 | 25 (69·4) | 0·9 | 36 | 16 (44·4) | 0·7 | 15 | 6 (40·0) | 1·0 |
| ≥4 | 99 | 68 (68·7) | ref | 99 | 40 (40·4) | ref | 38 | 15 (39·5) | ref |

**Abbreviations:** RT-PCR: reverse transcription polymerase chain reaction.
*P* values calculated by Chi-square test for binary explanatory variables and by logistic regression for explanatory variables with more than two levels, using the level with most observations as reference (ref). *P* values <0·05 marked in bold were considered statistically significant.
\* *P*-value for comparison of RT-PCR-positives and negatives, “not tested” omitted.

**Table 2:** Impact of index case characteristics on household secondary attack rate.

|  | n | Clinical symptoms<br>n (%) | P | n | Spike-specific IgG<br>n (%) | P | n | RT-PCR<br>n (%) | P |
| --- | --- | --- | --- | --- | --- | --- | --- | --- | --- |
| **Overall** | 179 |  |  | 179 |  |  | 70 |  |  |
| **Gender** |  |  |  |  |  |  |  |  |  |
| Male | 96 | 72 (75.0) | 0.4 | 96 | 41 (42.7) | 0.5 | 33 | 16 (48.5) | 0.7 |
| Female | 83 | 58 (69.9) |  | 83 | 40 (48.2) |  | 37 | 16 (43.2) |  |
| **Age, years** |  |  |  |  |  |  |  |  |  |
| <20 | 6 | 6 (100.0) | 1.0 | 6 | 2 (33.3) | 0.6 | 5 | 2 (40.0) | 0.9 |
| 20-59 | 157 | 109 (69.4) | ref | 157 | 68 (43.3) | ref | 59 | 25 (42.4) | ref |
| ≥60 | 16 | 15 (93.8) | 0.07 | 16 | 11 (68.8) | 0.06 | 6 | 5 (83.3) | 0.09 |
| **Fever** |  |  |  |  |  |  |  |  |  |
| Yes | 127 | 93 (73.2) | 0.8 | 127 | 67 (52.8) | **0.002** | 55 | 26 (47.3) | 0.6 |
| No | 52 | 37 (71.2) |  | 52 | 14 (26.9) |  | 15 | 6 (40.0) |  |
| **Cough** |  |  |  |  |  |  |  |  |  |
| Yes | 114 | 83 (72.8) | 0.9 | 114 | 52 (45.6) | 0.9 | 42 | 18 (42.9) | 0.6 |
| No | 65 | 47 (72.3) |  | 65 | 29 (44.6) |  | 28 | 14 (50.0) |  |
| **Dyspnoea** |  |  |  |  |  |  |  |  |  |
| Yes | 100 | 80 (80.0) | **0.01** | 100 | 53 (53.0) | **0.02** | 44 | 17 (38.6) | 0.1 |
| No | 79 | 50 (63.3) |  | 79 | 28 (35.4) |  | 26 | 15 (57.7) |  |

**Abbreviations:** RT-PCR: reverse transcription polymerase chain reaction.
*P* values calculated by Chi-square test for binary explanatory variables and by logistic regression for explanatory variables with more than two levels, using the level with most observations as reference (ref). *P* values <0.05 marked in bold were considered statistically significant.
<sup>a</sup>Each index case (n=112) may contribute to transmission to one or more household members.

**Figure 3.**
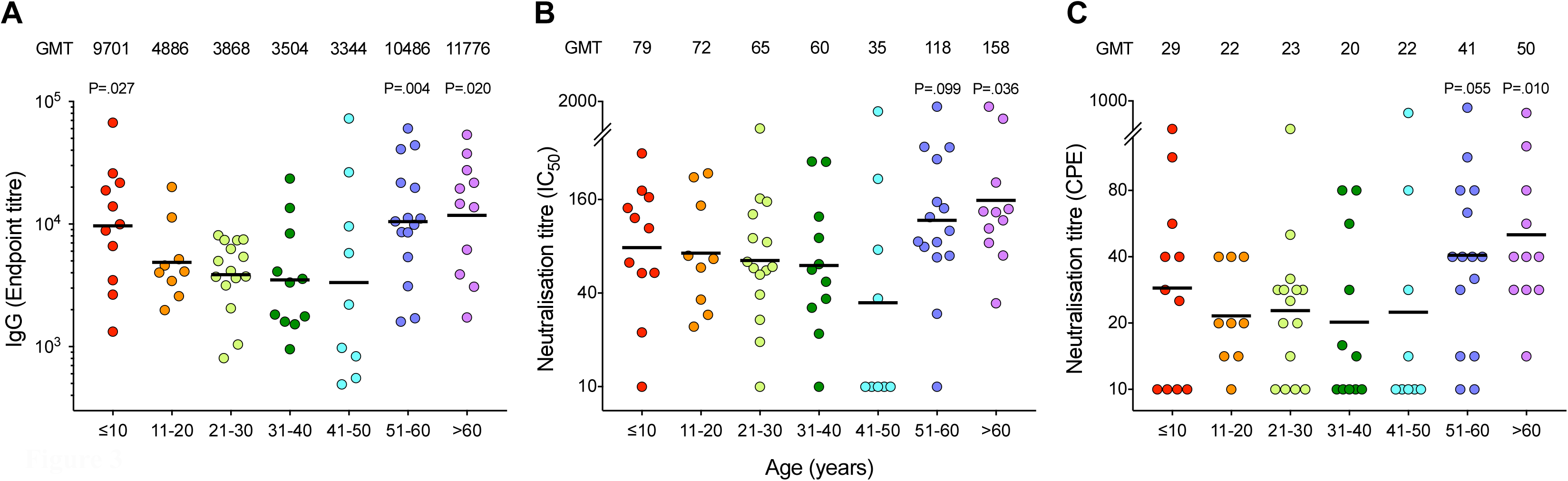
The SARS-CoV-2 antibody responses in seropositive household members. Sera from all household members were tested against the receptor-binding domain (RBD) of spike protein in screening ELISA. The positive samples (RBD IgG OD>0.5) were further tested for spike-specific IgG in confirmatory ELISA. Sera from all household members were tested in microneutralisation and virus neutralisation assays with live SARS-CoV-2/Human/NOR/Bergen1/2020 virus in a certified Biosafety Level 3 Laboratory. Household members with spike-specific IgG endpoint titre above 150 were defined as seropositive, and were divided into 10-year age cohorts, ≤10 years old n=11, 11-20 years old n=9, 21-30 years old n=15, 31-40 years old n=11, 41-50 years old n=9, 51-60 years old n=15, >60 years old n=11, in the age cohorts, respectively). Spike-specific IgG **(A)**, microneutralisation titres **(B)**, and virus neutralisation titres **(C)** are shown. The geometric mean titres (GMT) are shown above the graphs for each age cohort, and indicated by a horizontal line. Each symbol represents one subject. Mann-Whitney test was used in comparing antibody titres between age cohorts, 21-30 years as the reference group. *P*<0·05 were considered significant. All *P*<0·10 are noted. Two or more replicates were performed in all experiments. IC_50_, 50% inhibitory concentration. CPE, cytopathic effect.

### Comparison of RT-PCR and seroconversion

The seroconversion and RT-PCR positivity rates were further compared in the 70 household members who were RT-PCR tested during acute illness. We found that serological assays detected a higher number of infected household members than RT-PCR (44/70 vs 32/70), and thus are more sensitive than RT-PCR in detecting infected individuals (figure 4). Of the 32 household members who tested positive by RT-PCR during acute illness, only four (13%) did not seroconvert (table 1). If infection is defined by seroconversion or RT-PCR positivity, the overall attack rate was 47% among household members.

**Figure 4.**
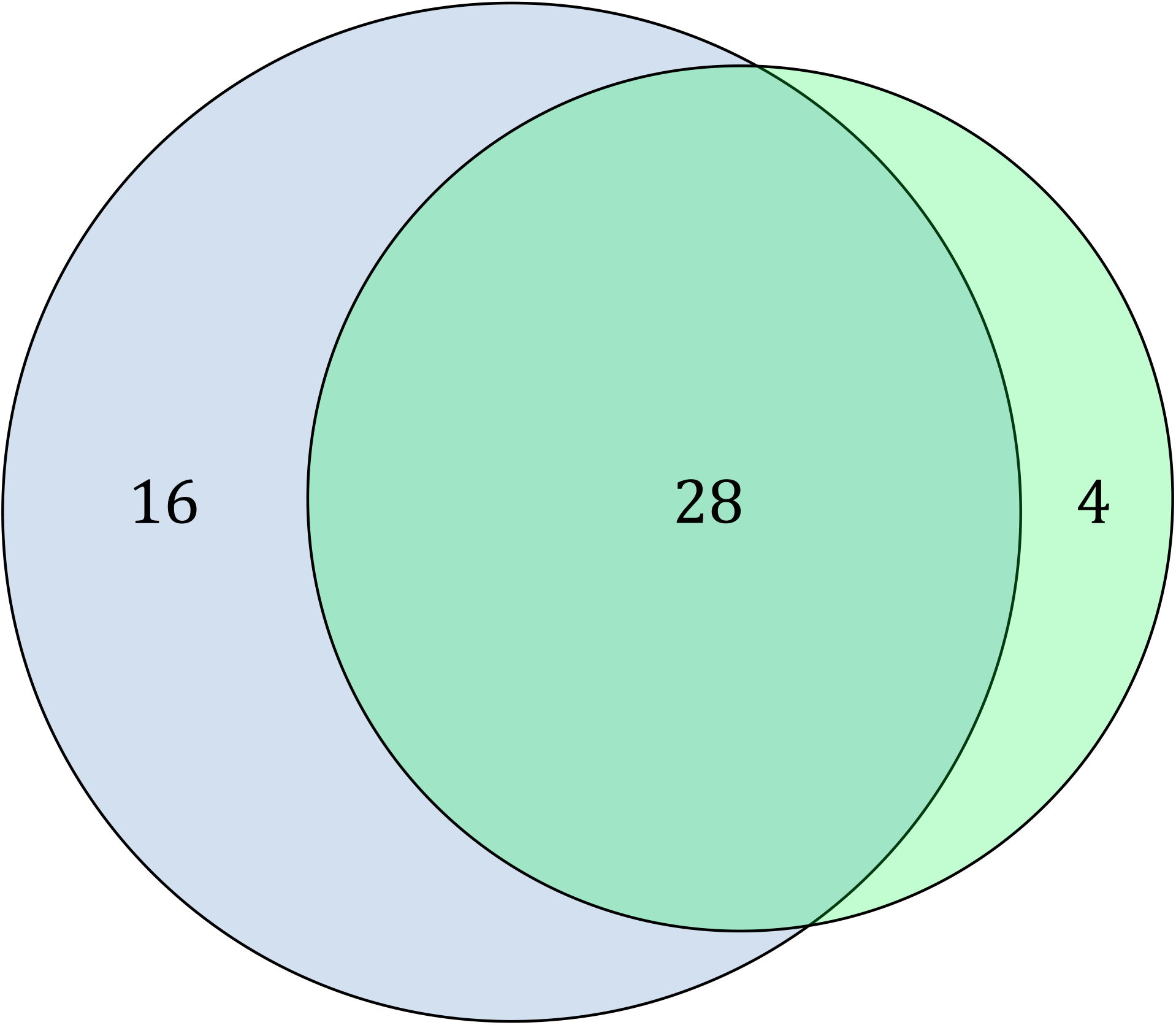
Serology detected more cases of infection in household members than RT-PCR. Venn-diagram of spike IgG positivity and RT-PCR positivity among 70 household members being tested by both methods. RT-PCR positives are indicated in light green and spike IgG positives in light blue. The overlapping region indicates household members positive by both methods. Twenty-two household members tested negative by both methods and are not shown in the figure.

As only symptomatic people were tested, RT-PCR positivity among asymptomatic household members was not assessed. Interestingly, among the household members who were RT-PCR tested, as many as 43% of the negatives seroconverted (table 1). Spike antibody titres were also compared between RT-PCR negative and RT-PCR positive members (figure 5D). Among RT-PCR tested household members who seroconverted, the spike antibody response was equally strong regardless of RT-PCR results.

**Figure 5.**
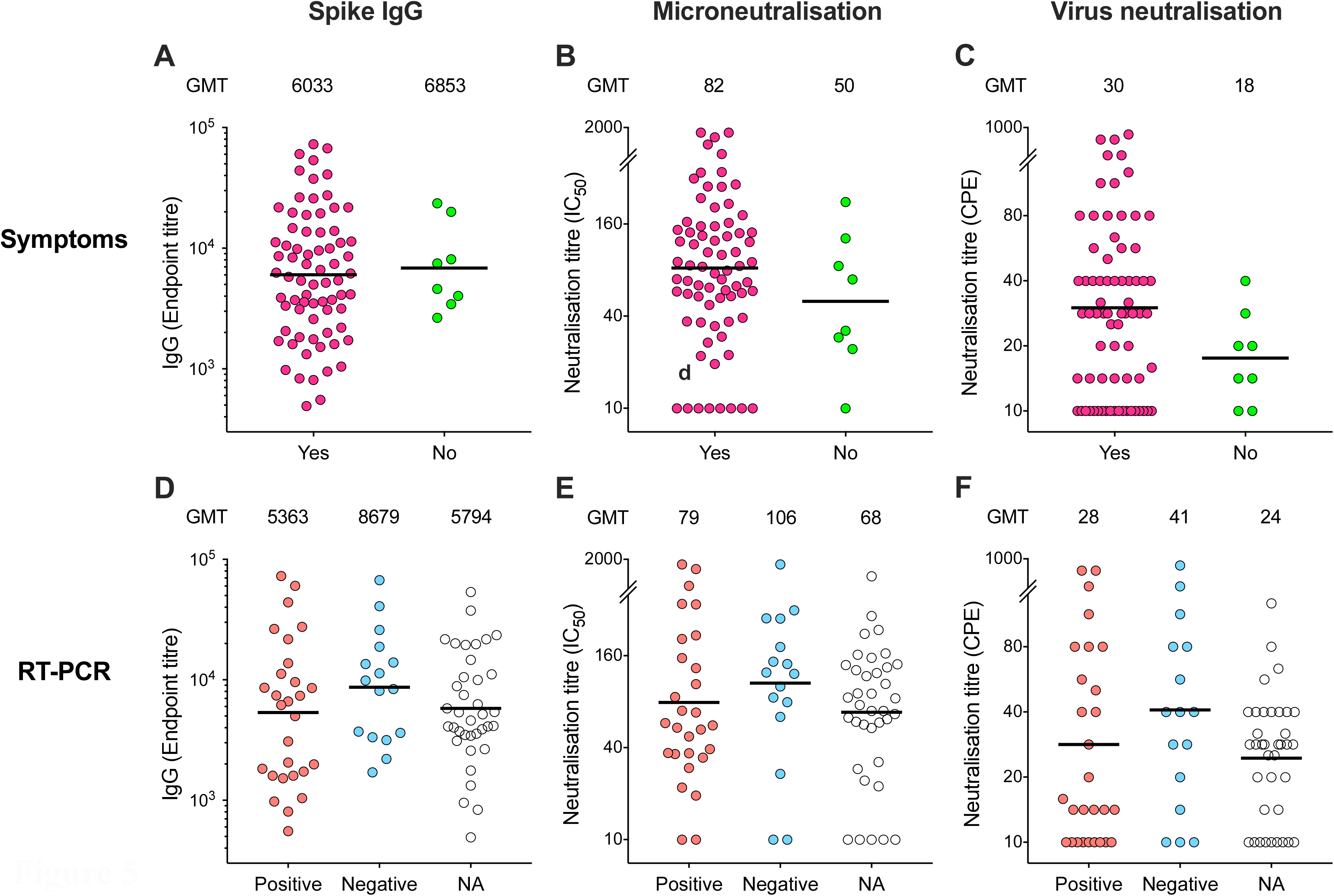
Serological responses compared to symptomatology and to SARS-CoV-2 RT-PCR results. Clinical symptoms of COVID-19 illness and SARS-CoV-2 RT-PCR results were collected from household members at the time of recruitment, blood samples were collected 6-8 weeks after diagnosis of the index case. Only symptomatic household members were tested by RT-PCR according to the testing algorithm at the centralized testing centre, therefore results are not available (NA) from all subjects. Spike-specific IgG **(A, D)**, microneutralisation **(B, E)** and virus neutralisation **(C, F)** titres from all seropositive household members are plotted against symptoms (n=73 in “Yes”, n=8 in “No” in **A-C**) and RT-PCR results (n=28 in “Positive”, n=16 for “Negative” and n=37 in “NA” in **D-F**). The geometric mean titres (GMT) for each subgroup are shown above the graphs, and indicated by a horizontal line. Mann-Whitney test was performed comparing symptomatic and asymptomatic subjects (in **A-C**) and RT-PCR positive and negative subjects (in **D-F**). No significant differences were found. Two or more replicates were performed in all experiments. IC_50_, 50% inhibitory concentration. CPE, cytopathic effect.

Interestingly, among children with symptoms compatible with COVID-19, only 11% (1/9) of those tested with RT-PCR were positive, while 60% (12/20) seroconverted. In contrast, among symptomatic persons ≥60 years, 89% (8/9) of those tested were RT-PCR positive, while 81% (13/16) seroconverted.

### Neutralising antibody responses

Neutralising antibodies can prevent reinfection with SARS-CoV-2 and we further analysed the neutralising antibody response by using the sensitive microneutralisation assay and the virus neutralisation assay to measure sterilising immunity. Significantly higher neutralisation titres were found in adults 60 years old (figure 3). Although children had higher spike-specific IgG than adults (21-30 years), they had similar titres of neutralising antibodies and some children did not have neutralising antibodies, particularly virus neutralising antibodies (figure 3). Interestingly, the levels of spike-specific antibodies and microneutralising antibodies were similar among symptomatic and asymptomatic individuals (figure 5A-B). There was a trend of higher virus neutralising antibodies among symptomatic household members, although not statistically significant (figure 5C). Among the household members who were RT-PCR tested, the levels of neutralising antibodies were equally high among RT-PCR positive and RT-PCR negative participants (figure 5E-F).

### Risk factors for transmission

The risk factors for household transmission are presented in table 2 and supplementary table 2. Household-members ≥60 years old (p=0·045), and those reporting symptoms (p<0·001) had higher risk of acquiring infection. Index-cases of seroconverting household-members had significantly higher titres of spike-specific IgG (p=0.044) and virus neutralisation (p=0.004) than index-cases of non-seroconverters. Whereas attack rates did not increase when the index case had a cough (p=0·9), transmission was more likely when the index cases had fever (p=0·002) or dyspnoea (p=0·02).

## Discussion

Studies on household transmission of SARS-CoV-2 provide crucial knowledge about the transmission dynamics of the virus in immunologically naïve individuals in a home environment characterized by limited personal protection. Norway contained community transmission at an early phase of the pandemic’s first wave by prompt implementation of measures to reduce contact between people in society. This ensured that our study was conducted with low levels of community transmission and negligible baseline immunity among the participants, which can otherwise confound household transmission studies. To investigate household transmission, we recruited the initial 112 households of RT-PCR confirmed SARS-CoV-2 index cases during the first month of the outbreak in Bergen, in a prospective, case-ascertained study. Our study was explicitly designed to measure household secondary attack rates based on the serological evaluation of SARS-CoV-2-specific antibodies. We found higher rates of transmission within households than previously reported (11-20), particularly in young children and elderly (>60 years old).

The overall household attack rate as measured by seroconversion among household members of RT-PCR confirmed, home-isolated cases was 45%. Currently, there are only two other studies that have estimated household attack rates based on seropositivity, 37·4% in Spain (7) and 35% in Brazil (22). Although both studies had large sample sizes in the early phase of the pandemic, they are population-based serosurveillance surveys which had high levels of community transmission and they cannot confirm that subjects were infected by a household member. Our study was specifically designed to assess household attack rates as measured by seropositivity in household members 6-8 weeks after onset of symptoms in the index case, with low prevalence of SARS-CoV-2 virus in the community. Thus, our data provide a more accurate estimate of attack rates.

We calculated attack rates based on SARS-CoV-2-specific antibodies in household members, whereas the majority of previous studies have ascertained transmission based on RT-PCR, with estimates of 7·6% to 38% (11-20). RT-PCR can only detect SARS-CoV-2 during the acute phase, and has been reported to have an unsatisfactory positivity rate (23, 24). Thus, the household secondary attack rates in RT-PCR-based studies is likely underestimated. This is supported by our finding that the seroconversion rate among RT-PCR negative household members(43%), was as high as the overall seropositivity rate among all household members (45%). Although we have a relatively small subgroup of RT-PCR negative household members during the inclusion period, our findings have two major implications. Firstly, using RT-PCR among household contacts of confirmed cases has a low predictive value, and solely relying on RT-PCR could consequently cause further transmission from false-negative cases to new individuals in both the household and the community. Secondly, our findings highlight that serological testing is equally or more effective than RT-PCR in confirming a final diagnosis of COVID-19, especially among household contacts. This is supported by several studies demonstrating the importance of serological testing to confirm cases (25, 26). However, among the household members in our study who had a positive RT-PCR test at the time of symptoms, 12% did not seroconvert. Extrapolation of this finding to our total cohort gives a total attack rate of 51%. Thus, the true attack rate in our study is likely higher than estimated solely by seroconversion. Our data also show that neutralisation assays seem to be less sensitive to detect attack rates than spike-specific IgG.

Children have been reported to be less affected by COVID-19 (27) and previous studies have reported a secondary attack rate of 4% to 23·1% among children (13, 14, 17). However, our study indicates that a larger proportion of children aged 0-12 years were infected (43%) in a household setting. The lower attack rates in previous studies may be due to the use of RT-PCR as a diagnostic method, consequently underestimating the number of secondary cases among children. Indeed, our data show a far lower positivity rate on RT-PCR among symptomatic children compared to serological testing, which contrasts with older age cohorts. Due to the low number of RT-PCR tested children in our study, this observation needs to be confirmed in larger studies, but suggests careful consideration of especially negative RT-PCR results in children. Age cut-offs as well as the testing criteria probably also play an important role in variations in reported SARS-CoV-2 infection risk in children. Estimates of transmission to children are also likely to be lower since children often present with milder symptoms, possibly resulting in lower testing rates. We conducted serological testing of all children among the household members, regardless of symptoms. With an attack rate above 40% as measured by seroconversion, our results show that children may have higher infection rates than has been previously reported (7, 13, 14, 17, 18, 28). The youngest index case was 15 years old in our study, and further studies of the role of children in transmission of the virus are urgently needed.

In our study, household members ≥60 years old appeared to have the highest attack rate (72%, table 1). Our study thus confirms findings from other studies on household transmission among older age groups (12, 15), while two studies have showed lower transmission to the oldest age cohort (13, 19). Moreover, we found neutralising antibodies in all but one of the seropositive participant aged ≥60 years (figure 3). As old age is strongly associated with morbidity and mortality, one may speculate that strong, neutralising antibody responses may be harmful.

The finding that index cases with fever and dyspnoea were more likely to transmit infection to others, is not surprising as patients with more severe symptoms may require closer follow-up and care, incurring increased risk of transmission. Similarly, the higher antibody-titres found in index cases of seroconverters may be a surrogate marker for higher and/or prolonged virus loads. It may appear counterintuitive that cough in the index case was not a significant risk factor for transmission. A likely explanation for this would be that, due to widespread awareness of this transmission route, cough would trigger household-members to use precautions such as distancing and mask use, while a person with other symptoms such as fever and dyspnoea may not be perceived as equally infectious.

According to a recent meta-analysis (5), an average of 15% of RT-PCR confirmed cases are asymptomatic. We found that 17% of asymptomatic household contacts seroconverted, and in addition, 43% of RT-PCR negative household members seroconverted. Thus, our findings show that close contacts of confirmed cases are potentially contagious, irrespective of being asymptomatic or having a negative RT-PCR result.

Whilst self-isolation of cases and good hygiene may prevent infection within a household, pre-existing immunity may also be important. Recently, pre-existing cross-reactive T-cell immunity derived from infection with human coronaviruses has been speculated to protect from infection (29). Although, the immune response to the SARS-CoV-2 virus is multifaceted and the correlates of protection from COVID-19 disease have yet to be defined. The presence of spike-specific antibodies does not directly correlate with protective immunity and therefore we used stringent serological assays measuring both microneutralising antibodies, which may prevent re-infection, and virus neutralising antibodies which provide sterilizing immunity. No neutralising antibodies were found in the household members who did not have spike-specific IgG (supplementary figure 3). When comparing the different assays, we found the highest attack rates measured by spike-specific antibodies, and lower numbers of household members developed neutralising antibodies. We did not specifically study the immunological mechanism of protection from infection household members who were not infected, and future studies on the impact of pre-existing cross-reactive immunity will be important in understanding the groups at highest risk for infection.

The strengths of our study are the centralized testing facility which allowed for the identification of all RT-PCR test positive cases in Bergen, the low levels of community spread and the stringent use of serological assays to define infected people, firstly by screening all subjects for RBD-specific antibodies, then confirming infection by SARS-CoV-2 spike ELISA. The study was specifically designed to identify household attack rate, and inclusion started with the first RT-PCR positive case in the city, followed by detailed interviews to differentiate between index cases and household members.

The interpretation of our findings has several limitations which may influence our estimation of attack rates. Despite high participation rates, there may have been a bias in who consented to participate, limiting and influencing our interpretation of results. There was also a risk that the index case was not correctly identified, although to minimize this, extensive telephone interviews were conducted once a positive case was identified. Likewise, we cannot exclude the possibility that some cases and household members had a common source of exposure outside the household, although the national shutdown appears to have eliminated the transmission of respiratory viruses in general, as evidenced by the abrupt drop in influenza cases. Lastly, the screening algorithm used by the municipality changed mid-March, with prioritisation of health-care workers and patients with underlying chronic conditions, potentially limiting the identification of patients.

In conclusion, we found a higher household attack rate of SARS-CoV-2 than previous studies, and show that serological testing is superior to RT-PCR-testing in assessing attack rates. Children are far more susceptible to household-transmission than previously reported, and relying on RT-PCR for diagnosis may miss the majority of infected children. This highlights the importance of including children when considering measures to reduce spread of SARS-CoV-2 virus. The risk of transmission was highest from index cases with dyspnoea, fever and high titres of neutralising antibodies, all potential surrogate markers for severity of disease, illustrating that transmission risk increases with the need for close care.

## Data Availability

The data is available from the corresponding author upon reasonable request

## Author Contributions

NL, BB, RJC, KAB, KGIM, and CT designed the study. KK and DL recruited the participants. SL, KAB, MCT, and AM conducted laboratory analysis and developed the assays. FZ developed and ran all the neutralisation assays. CT recruited all the children from households. FK developed the two step ELISA. KK, FZ, BB, RJC, and NL analysed the data and wrote the manuscript. All authors have read and approved the final version of the manuscript.

## Supplementary material

### Supplementary methods

#### SARS-COV-2 RBD and spike protein production

SARS-COV-2 RBD and spike stabilized trimer constructs have been described by Stadlbauer and colleagues (6). We expressed and purified the RBD as previously described (6). After clarifying cell culture medium by centrifugation, the supernatant was loaded into a HiFliQ Ni Advance column (ProteinArk), and we determined protein concentration with Quick Start Bradford Protein Assay using BSA as standards (Bio-Rad). Initially, we prepared spike trimer according to Staldbauer et al. 2020 with the exception that we captured spike trimer as above. We gradually shifted for expression in ExpiCHO cell following manufacturers protocols and recommendations (ThermoFisher Scientific) in Optimum Growth flask (Thomson Instrument Company). In later batches, we found that extended expression time in 32°C ExpiCHO or Expi293F cells gave higher yields and less lower molecular weight contaminations in Corning Erlenmeyer or Optimum Growth flasks (Esposito et al. 2020, Herrera et al. 2020; PMID: 32504802; preprint PMID: 32587972).

#### Enzyme-linked immunosorbent assay (ELISA)

The ELISA for detecting SARS-CoV-2 RBD-specific antibodies was performed as previously described (6) and used to screen all index case and household members serum samples for RBD-specific antibodies. ELISA plates (Thermo Fisher) were coated with 50 μL of SARS-CoV-2 RBD protein (2 μg/mL) in PBS at 4°C overnight and sera tested at 1:100 dilution, followed by detection with horseradish peroxidase (HRP) labeled goat anti-human IgG (Sigma) and development with 3,3’,5,5’-tetramethylbenzidine (TMB) (BDbiosciences, E-2886) for 10 minutes. Plates were read at 450 nm and 620 nm (background) using a synergy H1 plate reader (BioTek). A negative cut-off OD <0·5036 was defined by using historical serum samples collected before 2020 (n=128). A confirmation spike ELISA was performed to quantify the SARS-CoV-2 spike-specific antibodies in subjects with positive screening results, as described for the screening ELISA, with the following modification. Plates were coated with 2 μg/mL of spike protein. Serum samples were 5-fold serially diluted from 1:100. Plates were incubated for 15 minutes with TMB before stopping. The endpoint titres were calculated using Prism 8. Endpoint titre was defined as the highest serum dilution to give an OD value ≥ 3 standard deviations above the historical serum samples collected before the pandemic (n = 128). Individuals with undetectable levels of antibodies were assigned an endpoint titre of 50 for plotting and calculation purposes. Historical serum samples collected before 2019 were defined as seronegative in the RBD ELISA, which was confirmed with RT-PCR negative healthcare worker sera collected during the pandemic (unpublished results). As recruitment was initiated with the very first case of SARS-CoV-2 identified in Bergen municipality, we assume all participants were assumed seronegative at baseline and seropositivity with spike-specific IgG is termed seroconversion.

#### SARS-CoV-2 Virus

A clinical SARS-CoV-2 strain was isolated in-house from a throat swab of a Norwegian RT-PCR confirmed patient, propagated in Vero cells, and named as SARS-CoV-2/Human/NOR/Bergen1/2020.

#### Microneutralisation assay

The microneutralisation (MN) assay was used to investigate the microneutralising titres, which can prevent infection. The assay was performed in a certified Biosafety Level 3 Laboratory. Serum samples were tested against clinical isolated virus: SARS-CoV-2/Human/NOR/Bergen1/2020 (GISAID accession number pending). Two or more biological replicates were performed. Briefly, serum samples were heat inactivated at 56°C for 60 minutes, analysed in serial dilutions (duplicated, starting from 1:20), and mixed with 100 TCID50 viruses in 96-well plates and incubated for one hour at 37 °C. Mixtures were transferred to 96-well plates seeded with Vero cells. The plates were incubated at 37 °C for 24 hours. Cells were fixed and permeabilized with methanol and 0.6% H2O2, and incubated with rabbit monoclonal IgG against SARS-CoV2 NP (Sino Biological). Cells were further incubated with biotinylated goat anti-rabbit IgG (H+L) (Southern Biotech), and Extravidin-peroxidase (Sigma-Aldrich). The reactions were developed with o-phenylenediamine dihydrochloridec (OPD) (Sigma-Aldrich). The MN titre was determined as the reciprocal of the serum dilution giving 50% inhibition of virus infectivity. Negative titres (<20) were assigned a value of 10 for calculation purposes.

#### Virus neutralisation assay

The virus neutralisation (VN) assay was performed in a certified Biosafety Level 3 facility. Serum samples were tested against clinical isolated virus: SARS-CoV-2/Human/NOR/Bergen1/2020. Two or more biological replicates were performed. Briefly, serum samples were heat-inactivated at 56°C for 60 min, analysed in serial dilutions (duplicate, starting from 1:20), and mixed with 100 TCID50 viruses in 96-well plates and incubated for one hour at 37°C. Mixtures were transferred to 96-well plates with Vero cells. The plates were incubated at 37°C for 4-5 days. All wells were examined under microscope for cytopathic effect (CPE). The VN titre was determined as the reciprocal of the highest serum dilution giving no CPE. Negative titres (<20) were assigned a value of 10 for calculation purpose.

**Supplementary figure 1.**
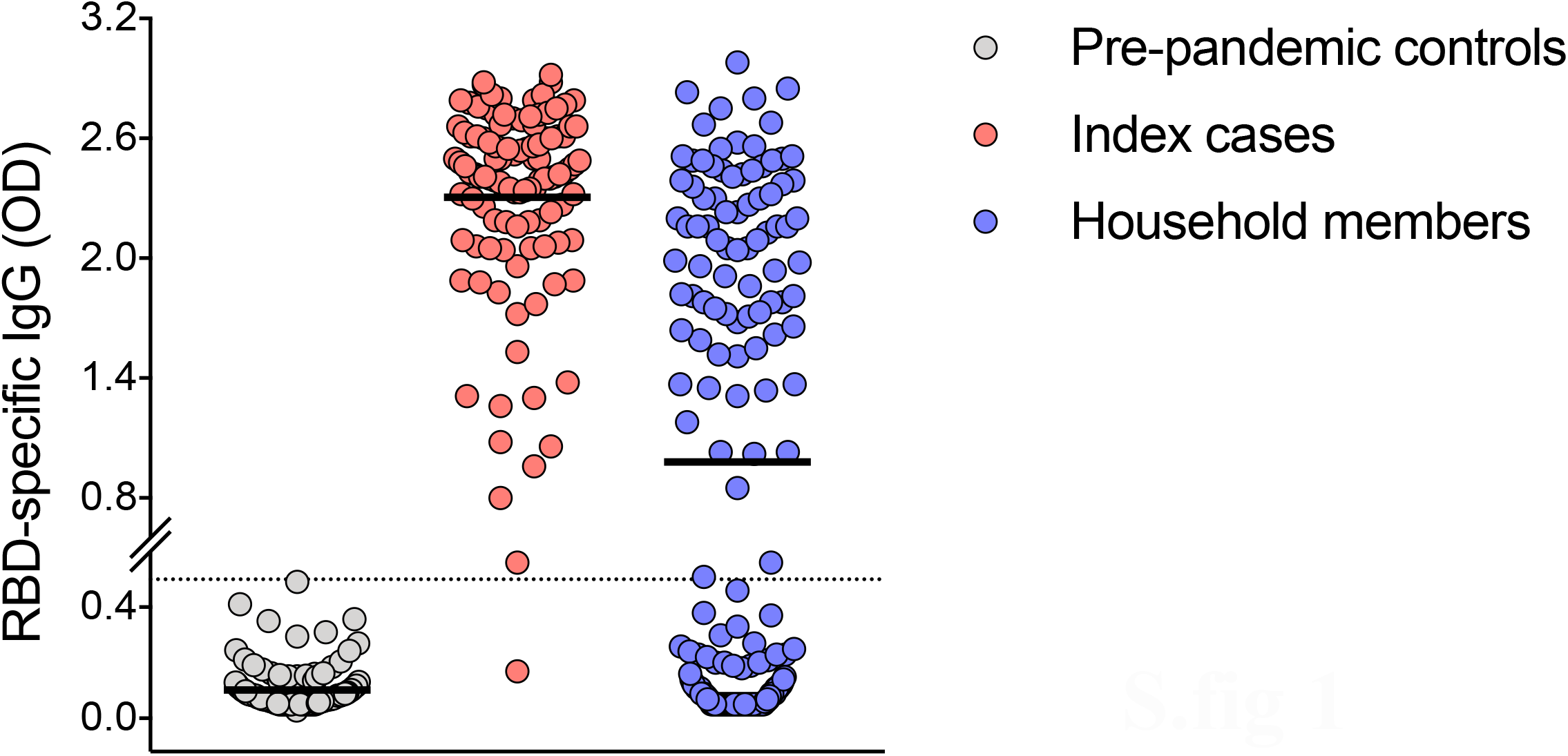
Screening receptor-binding domain IgG from pre-pandemic controls, index cases and household members. A panel of pre-pandemic sera (n=128) from blood donors was established from historic samples biobanked before 2019. All 128 pre-pandemic controls were screened against the receptor-binding domain (RBD) of spike protein of SARS-CoV-2, using the same conditions as for all index cases and household members. A positive cut-off of RBD screening ELISA was defined as three standard deviations above the mean of 128 pre-pandemic controls. All index cases and household members with RBD IgG optical density (OD)>0·5 were further measured for anti-spike IgG. RBD-specific IgG are shown as the optical density at 1/100 dilution of sera. Each symbol represents one subject. The mean of OD in each group is marked as a horizontal line. The dotted line indicates OD 0·5 as the cut-off. Duplicates were performed in ELISA.

**Supplementary figure 2.**
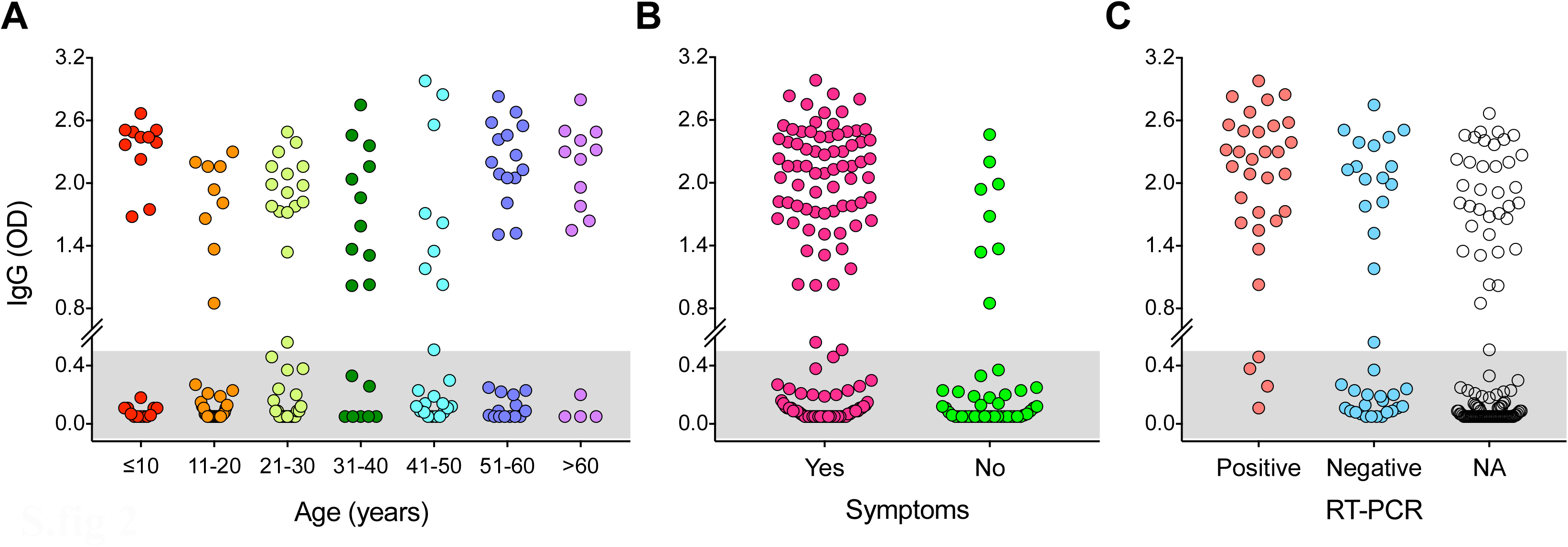
Screening receptor-binding domain IgG from all household members by age-cohorts, symptoms and RT-PCR results. All household members sera were screened by receptor-binding domain ELISA at a dilution of 1/100. The gray shaded area indicates the negative samples below the cut-off (optical density (OD) >0·5) defined by historical blood donor samples. Household members are divided by 10-year age cohorts **(A)**, clinical symptoms of COVID-19 illness **(B)** and SARS-CoV-2 RT-PCR results **(C)**. Each symbol represents one subject. NA indicates not available because the household member was not tested by RT-PCR. Duplicates were performed in all experiments.

**Supplementary figure 3.**
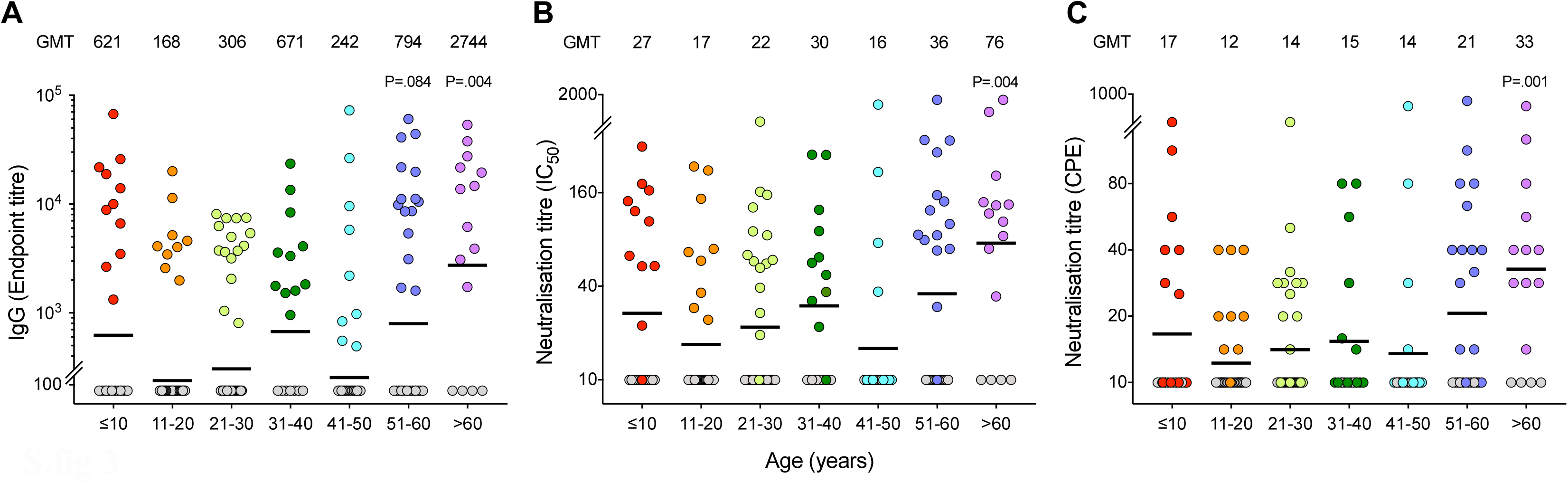
The SARS-CoV-2 antibody responses in all 179 household members. Sera from all household members were tested against the receptor-binding domain (RBD) of spike protein in screening ELISA. The positive samples (RBD IgG OD>0·5) were further tested for spike-specific IgG in confirmatory ELISA. Sera from all household members were tested in microneutralisation and virus neutralisation assays with live SARS-CoV-2/Human/NOR/Bergen1/2020 virus in a certified Biosafety Level 3 Laboratory. Household members, regardless of their spike-specific IgG endpoint titre, were divided into 10-year age cohorts (≤10 years old n=23, 11-20 years old n=34, 21-30 years old n=36, 31-40 years old n=18, 41-50 years old n=24, 51-60 years old n=29, >60 years old n=15). Spike-specific IgG **(A)**, microneutralisation titres **(B)**, and virus neutralisation titres **(C)** are shown. The geometric mean titres (GMT) are noted above the graphs for each age cohort, and indicated by a horizontal line. Each symbol represents one subject. Seropositive subjects are shown in colour and seronegative ones are indicated by gray symbols. Mann-Whitney test was used in comparing antibody titres between age cohorts, 21-30 years as the reference group. *P*<0·05 were considered significant. All *P*<0.50 are noted. Two or more replicates were performed in all experiments. IC_50_, 50% inhibitory concentration. CPE, cytopathic effect.

**Supplementary table 1:** Demographic, clinical and serological characteristics by index cases and household members.

**S.fig 3**
|  | Index cases<br>n (%) | Household members<br>n (%) | *P* |
| --- | --- | --- | --- |
| **Overall** | 112 | 179 |  |
| **Sex** |  |  |  |
| Male | 54 (48) | 80 (45) | 0·6 |
| Female | 58 (52) | 99 (55) |  |
| **Age, years** |  |  |  |
| Mean (SD) | 44 (15) | 33 (19) | **<0·001** |
| Median (Q1,Q3) | 45 (31,54) | 30 (17, 50) |  |
| **Clinical symptoms** |  |  |  |
| Yes | 112 (100) | 130 (73) | **<0·001** |
| No | 0 (0) | 49 (27) |  |
| **RT-PCR** |  |  |  |
| Positive | 112 (100) | 32 (18) | **<0·001** |
| Negative | 0 (0) | 38 (21) |  |
| Not tested | 0 (0) | 109 (61) |  |
| **Serology** |  |  |  |
| Spike-specific antibodies | 111 (99) | 81 (45) | **<0·001** |
| Microneutralising antibodies | 100 (89) | 71 (40) | **<0·001** |
| Virus neutralising antibodies | 65 (58) | 51 (28) | **<0·001** |

**Abbreviations:** Q1: 25<sup>th</sup> percentile; Q3: 75<sup>th</sup> percentile; RT-PCR: reverse transcription polymerase chain reaction.
*P* values calculated by Chi-square test for proportions and Mann-Whitney test for age. *P* values <0·05 marked in bold were considered statistically significant.

**Supplementary table 2:** Household secondary attack rate by characteristics of household members, index cases and household size.

|  | n | Clinical symptoms<br>n (%) | P | Spike-specific<br>IgG<br>n (%) | P | MN<br>n (%) | P | VN<br>n (%) | P | n | RT-PCR<br>n (%) | P |
| --- | --- | --- | --- | --- | --- | --- | --- | --- | --- | --- | --- | --- |
|  | 179 | 130 (72.6) |  | 81 (45.3) |  | 70 (39.1) |  | 51 (28.5) |  | 70 | 32 (45.7) |  |
| Household member characteristics |  |  |  |  |  |  |  |  |  |  |  |  |
| Sex |  |  |  |  |  |  |  |  |  |  |  |  |
| Male | 80 | 57 (71.3) | 0.7 | 38 (47.5) | 0.6 | 33 (41.2) | 0.6 | 24 (30.0) | 0.7 | 34 | 15 (44.1) | 0.8 |
| Female | 99 | 73 (73.7) |  | 43 (43.4) |  | 37 (37.4) |  | 27 (27.3) |  | 17 | 17 (47.2) |  |
| Age, years |  |  |  |  |  |  |  |  |  |  |  |  |
| 0-10 | 23 | 17 (73.9) | 0.6 | 11 (47.8) | 0.6 | 10 (43.5) | 0.6 | 7 (30.4) | 0.8 | 7 | 1 (14.3) | 0.2 |
| 11-20 | 34 | 22 (64.7) | 0.9 | 9 (26.5) | 0.2 | 9 (26.4) | 0.4 | 6 (17.6) | 0.3 | 9 | 2 (22.2) | 0.3 |
| 21-30 | 36 | 24 (66.7) | ref | 15 (41.7) | ref | 13 (36.1) | ref | 10 (27.8) | ref | 19 | 8 (42.1) | ref |
| 31-40 | 18 | 14 (77.8) | 0.4 | 11 (61.1) | 0.2 | 10 (55.6) | 0.2 | 4 (22.2) | 0.7 | 8 | 4 (50.0) | 0.7 |
| 41-50 | 24 | 20 (83.3) | 0.2 | 9 (37.5) | 0.7 | 4 (16.7) | 0.1 | 3 (12.5) | 0.2 | 9 | 5 (55.6) | 0.5 |
| 51-60 | 29 | 19 (65.5) | 0.9 | 15 (51.7) | 0.4 | 13 (44.8) | 0.5 | 11 (37.9) | 0.4 | 11 | 6 (54.5) | 0.5 |
| >60 | 15 | 14 (93.3) | 0.08 | 11 (73.3) | 0.046 | 11 (73.3) | 0.02 | 10 (66.7) | 0.01 | 7 | 6 (85.7) | 0.07 |
| Symptoms |  |  |  |  |  |  |  |  |  |  |  |  |
| Yes | 130 | .. | .. | 73 (56.2) | <0.001 | 63 (48.5) | <0.001 | 47 (36.2) | <0.001 | 66 | 32 (48.5) | 0.06 |
| No | 49 | .. |  | 8 (16.3) |  | 7 (14.3) |  | 4 (8.2) |  | 4 | 0 (0.0) |  |
| RT-PCR results |  |  |  |  |  |  |  |  |  |  |  |  |
| Positive | 32 | 32 (100) | 0.06 | 28 (87.5) | <0.001 | 25 (78.1) | <0.001 | 13 (40.6) | 0.4 |  | .. | .. |
| Negative | 38 | 34 (89.5) |  | 16 (42.1) |  | 14 (36.8) |  | 12 (31.6) |  |  | .. |  |
| Not tested | 109 | 64 (58.7) |  | 37 (33.9) |  | 31 (28.4) |  | 26 (23.9) |  |  | .. |  |
| Household size |  |  |  |  |  |  |  |  |  |  |  |  |
| 2 | 44 | 37 (84.1) | 0.06 | 25 (56.8) | 0.07 | 22 (50.0) | 0.08 | 19 (43.2) | 0.01 | 17 | 11 (64.7) | 0.09 |
| 3 | 36 | 25 (69.4) | 0.9 | 16 (44.4) | 0.7 | 14 (38.9) | 0.6 | 10 (27.8) | 0.5 | 6 | 6 (40.0) | 1.0 |
| ≥4 | 99 | 68 (68.7) | ref | 40 (40.4) | ref | 34 (34.3) | ref | 22 (22.2) | ref | 15 | 15 (39.5) | Ref |
| Index characteristics<sup>a</sup> |  |  |  |  |  |  |  |  |  |  |  |  |
| Gender |  |  |  |  |  |  |  |  |  |  |  |  |
| Male | 96 | 72 (75.0) | 0.4 | 41 (42.7) | 0.5 | 34 (35.4) | 0.3 | 26 (27.1) | 0.7 | 33 | 16 (48.5) | 0.7 |
| Female | 83 | 58 (69.9) |  | 40 (48.2) |  | 36 (43.4) |  | 25 (30.1) |  | 37 | 16 (43.2) |  |
| Age, years |  |  |  |  |  |  |  |  |  |  |  |  |
| <20 | 6 | 6 (100.0) | 1.0 | 2 (33.3) | 0.6 | 1 (16.7) | 0.3 | 1 (16.7) | 0.6 | 5 | 2 (40.0) | 0.9 |
| 20-59 | 157 | 109 (69.4) | ref | 68 (43.3) | ref | 58 (36.9) | ref | 41 (26.1) | ref | 59 | 25 (42.3) | ref |
| ≥60 | 16 | 15 (93.8) | 0.07 | 11 (68.8) | 0.06 | 11 (68.8) | 0.02 | 9 (56.2) | 0.02 | 6 | 5 (83.3) | 0.09 |
| BMI, kg/m<sup>2</sup> |  |  |  |  |  |  |  |  |  |  |  |  |
| Underweight (<18.5) | 5 | 5 (100.0) | 1.0 | 2 (40.0) | 0.7 | 1 (20.0) | 0.3 | 1 (20.0) | 0.6 | 4 | 1 (25.0) | 0.8 |
| Normal (18.5-24.9) | 101 | 73 (72.3) | ref | 49 (48.5) | ref | 43 (42.6) | ref | 31 (30.7) | ref | 46 | 22 (47.8) | ref |
| Overweight (25.0-29.9) | 57 | 39 (68.4) | 0.6 | 22 (38.6) | 0.2 | 20 (35.1) | 0.4 | 14 (24.6) | 0.4 | 14 | 6 (42.9) | 0.7 |
| Obese (≥30.0) | 15 | 12 (80.0) | 0.5 | 8 (53.3) | 0.7 | 6 (40.0) | 0.9 | 5 (33.3) | 0.8 | 6 | 3 (50.0) | 0.9 |
| Clinical symptoms |  |  |  |  |  |  |  |  |  |  |  |  |
| Sick < 14 days |  |  |  |  |  |  |  |  |  |  |  |  |
| Yes | 94 | 67 (71·3) | 0·4 | 42 (44·7) | 0·6 | 35 (37·2) | 0·5 | 27 (28·7) | 0·9 | 35 | 17 (48·6) | 0·7 |
| No | 65 | 50 (76·9) |  | 32 (49·2) |  | 27 (41·5) |  | 18 (27·7) |  | 27 | 12 (44·4) |  |
| NA | 20 | .. |  | .. |  | .. |  | .. |  | 8 | .. |  |
| Cough |  |  |  |  |  |  |  |  |  |  |  |  |
| Yes | 114 | 83 (72·8) | 0·9 | 52 (45·6) | 0·9 | 46 (40·4%) | 0·7 | 33 (28·9) | 0·9 | 42 | 18 (42·9) | 0·6 |
| No | 65 | 47 (72·3) |  | 29 (44·6) |  | 24 (36·9) |  | 18 (27·7) |  | 28 | 14 (50·0) |  |
| Fever |  |  |  |  |  |  |  |  |  |  |  |  |
| Yes | 127 | 93 (73·2) | 0·8 | 67 (52·8) | **0·002** | 58 (45·7) | **0·005** | 44 (34·6) | **0·004** | 55 | 26 (47·3) | 0·6 |
| No | 52 | 37 (71·2) |  | 14 (26·9) |  | 12 (23·1) |  | 7 (13·5) |  | 15 | 6 (40·0) |  |
| Dyspnoea |  |  |  |  |  |  |  |  |  |  |  |  |
| Yes | 100 | 80 (80·0) | **0·01** | 53 (53·0) | **0·02** | 46 (46·0) | **0·03** | 35 (35·0) | **0·03** | 44 | 17 (38·6) | 0·1 |
| No | 79 | 50 (63·3) |  | 28 (35·4) |  | 24 (30·4) |  | 16 (20·3) |  | 26 | 15 (57·7) |  |
| Fatigue |  |  |  |  |  |  |  |  |  |  |  |  |
| Yes | 164 | 118 (72·0) | 0·5 | 74 (45·1) | 0·9 | 64 (39·0) | 0·9 | 47 (28·7) | 0·9 | 64 | 29 (45·3) | 0·8 |
| No | 15 | 12 (80·0) |  | 7 (46·7) |  | 6 (40·0) |  | 4 (26·7) |  | 6 | 3 (50·0) |  |
| Myalgia |  |  |  |  |  |  |  |  |  |  |  |  |
| Yes | 120 | 91 (75·8) | 0·2 | 58 (48·3) | 0·2 | 52 (43·3) | 0·1 | 36 (30·0) | 0·5 | 51 | 21 (41·2) | 0·2 |
| No | 59 | 39 (66·1) |  | 23 (39·0) |  | 18 (30·5) |  | 15 (25·4) |  | 19 | 11 (57·9) |  |
| Headache |  |  |  |  |  |  |  |  |  |  |  |  |
| Yes | 133 | 102 (76·7) | **0·04** | 61 (45·9) | 0·8 | 51 (38·3) | 0·7 | 39 (29·3) | 0·7 | 54 | 23 (42·6) | 0·3 |
| No | 46 | 28 (60·9) |  | 20 (43·5) |  | 19 (41·3) |  | 12 (26·1) |  | 16 | 9 (56·2) |  |
| **Comorbidities<sup>b</sup>** |  |  |  |  |  |  |  |  |  |  |  |  |
| Any comorbidities |  |  |  |  |  |  |  |  |  |  |  |  |
| Yes | 65 | 50 (76·9) | 0·3 | 34 (52·3) | 0·2 | 32 (49·2) | **0·04** | 23 (35·4) | 0·1 | 26 | 14 (53·8) | 0·3 |
| No | 114 | 80 (70·2) |  | 47 (41·2) |  | 38 (33·3) |  | 28 (24·6) |  | 44 | 18 (40·9) |  |
| DM type 1 or 2 |  |  |  |  |  |  |  |  |  |  |  |  |
| Yes | 3 | 2 (66·7) | 0·8 | 2 (66·7) | 0·5 | 2 (66·7) | 0·3 | 1 (33·3) | 0·9 | 1 | 1 (100·0) | 0·3 |
| No | 176 | 128 (72·7) |  | 79 (44·9) |  | 68 (38·6) |  | 50 (28·4) |  | 69 | 31 (44·9) |  |
| Asthma |  |  |  |  |  |  |  |  |  |  |  |  |
| Yes | 17 | 10 (58·8) | 0·2 | 11 (64·7) | 0·09 | 10 (58·8) | 0·08 | 5 (29·4) | 0·9 | 5 | 3 (60·0) | 0·5 |
| No | 162 | 120 (74·1) |  | 70 (43·2) |  | 60 (37·0) |  | 46 (28·4) |  | 65 | 29 (44·6) |  |
| CVD |  |  |  |  |  |  |  |  |  |  |  |  |
| Yes | 5 | 4 (80·0) | 0·7 | 4 (80·0) | 0·1 | 4 (80·0) | 0·06 | 4 (80·0) | **0·01** | 1 | 1 (100·0) | 0·3 |
| No | 174 | 126 (72·4) |  | 77 (44·3) |  | 66 (37·9) |  | 47 (27·0) |  | 69 | 31 (44·9) |  |
| Hypertension |  |  |  |  |  |  |  |  |  |  |  |  |
| Yes | 14 | 9 (64·3) | 0·5 | 6 (42·9) | 0·9 | 5 (35·7) | 0·8 | 5 (35·7) | 0·5 | 3 | 1 (33·3) | 0·7 |
| No | 165 | 121 (73·3) |  | 75 (45·5) |  | 65 (39·4) |  | 46 (27·9) |  | 67 | 31 (46·3) |  |
| Rheumatic disease |  |  |  |  |  |  |  |  |  |  |  |  |
| Yes | 9 | 4 (44·4) | **0·052** | 5 (55·6) | 0·5 | 3 (33·3) | 0·7 | 2 (22·2) | 0·7 | 3 | 1 (33·3) | 0·7 |
| No | 170 | 126 (74·1) |  | 76 (44·7) |  | 67 (39·4) |  | 49 (28·8) |  | 67 | 31 (46·3) |  |
| Immunodeficiency<sup>c</sup> |  |  |  |  |  |  |  |  |  |  |  |  |
| Yes | 5 | 4 (80·0) | 0·7 | 4 (80·0) | 0·1 | 4 (80·0) | 0·06 | 2 (40·0) | 0·6 | 4 | 2 (50·0) | 0·9 |
| No | 174 | 126 (72·4) |  | 77 (44·3) |  | 66 (37·9) |  | 49 (28·2) |  | 66 | 30 (45·5) |  |
| **Medication** |  |  |  |  |  |  |  |  |  |  |  |  |
| Inhaled corticosteroid |  |  |  |  |  |  |  |  |  |  |  |  |
| Yes | 12 | 6 (50·0) | **0·07** | 5 (41·7) | 0·8 | 3 (25·0) | 0·3 | 2 (16·7) | 0·3 | 3 | 1 (33·3) | 0·7 |
| No | 167 | 43 (25·7) |  | 76 (45·5) |  | 67 (40·1) |  | 49 (29·3) |  | 67 | 31 (46·3) |  |
| Oral corticosteroids |  |  |  |  |  |  |  |  |  |  |  |  |
| Yes | 2 | 1 (50·0) | 0·5 | 2 (100·0) | 0·2 | 2 (100·0) | 0·2 | 0 (0·0) | 1 | 1 | 1 (100·0) | 0·5 |
| No | 177 | 129 (72.9) |  | 79 (44.6) |  | 68 (38.4) |  | 51 (28.8) |  | 69 | 31 (44.9) |  |
| **Smoking** |  |  |  |  |  |  |  |  |  |  |  |  |
| Yes | 5 | 4 (80.0) | 0.8 | 3 (60.0) | 0.4 | 2 (40.0) | 0.9 | 1 (20.0) | 0.8 | 1 | 1 (100.0) | 1.0 |
| Smoked earlier | 56 | 39 (69.6) | 0.6 | 28 (50.0) | 0.3 | 25 (44.6) | 0.3 | 19 (33.9) | 0.3 | 16 | 7 (43.8) | 0.9 |
| Never smoked | 118 | 87 (73.7) | ref | 50 (42.4) | ref | 43 (36.4) | ref | 31 (26.3) | ref | 53 | 24 (45.3) | ref |

**Abbreviations:** CVD: cardiovascular disease; MN: microneutralising antibodies; NA: not available; RT-PCR: reverse transcription polymerase chain reaction; VN: virus neutralising antibodies.
<sup>a</sup>Each index case (n=112) may contribute to transmission to one or more household members (n=179).
<sup>b</sup>Comorbidities: no index cases reported having HIV, infection, active malignancy, chronic renal disease, chronic hepatic disease and dementia. Analysis is not reported for rarely reported comorbidities (chronic lung disease (n=1) and rarely used medications (angiotensin-converting enzyme inhibitors (n=1)).
<sup>c</sup>Immunodeficiency: primary immunodeficiency, human immunodeficiency virus, organ transplant, bone marrow transplant, chemotherapy and other immunosuppressive treatment (including disease-modifying anti-rheumatic drugs and biological therapy).
*P* values calculated by Chi-square test for binary explanatory variables and by logistic regression for explanatory variables with more than two levels, using the level with most observations as reference (ref). *P* values < 0.05 marked in bold were considered statistically significant.

## Notes

### Competing Interest Statement

The authors have declared no competing interest.

### Author Declarations

Western Norway Local Ethics Committee (#118664).

## References

1. Organization WH. WHO Coronavirus Disease (COVID-19) Dashboard 2020 [updated 2nd September 2020. Available from: https://covid19.who.int/.

2. Folkehelseinstituttet. En person har testet positivt på koronavirus 2020 [Available from: https://www.fhi.no/nyheter/2020/en-person-har-testet-positivt-pa-koronavirus/.

3. Regjeringen.no. Omfattende tiltak for å bekjempe koronaviruset 2020 [updated 12th March 2020.

4. Folkehelseinstituttet. COVID-19 Ukerapport - uke 35 [Available from: https://www.fhi.no/contentassets/8a971e7b0a3c4a06bdbf381ab52e6157/vedlegg/andre-halvar-2020/2020-09-02-ukerapport-uke-35-covid-19.pdf.

5. Byambasuren O, Cardona M, Bell K, Clark J, McLaws M-L, Glasziou P. Estimating the extent of asymptomatic COVID-19 and its potential for community transmission: systematic review and meta-analysis. medRxiv. 2020:2020.05.10.20097543.

6. Stadlbauer D, Amanat F, Chromikova V, Jiang K, Strohmeier S, Arunkumar GA, et al. SARS-CoV-2 Seroconversion in Humans: A Detailed Protocol for a Serological Assay, Antigen Production, and Test Setup. Curr Protoc Microbiol. 2020;57(1):e100.

7. Pollan M, Perez-Gomez B, Pastor-Barriuso R, Oteo J, Hernan MA, Perez-Olmeda M, et al. Prevalence of SARS-CoV-2 in Spain (ENE-COVID): a nationwide, population-based seroepidemiological study. Lancet. 2020;396(10250):535–44.

8. Zhao J, Yuan Q, Wang H, Liu W, Liao X, Su Y, et al. Antibody responses to SARS-CoV-2 in patients of novel coronavirus disease 2019. Clin Infect Dis. 2020.

9. Gudbjartsson DF, Norddahl GL, Melsted P, Gunnarsdottir K, Holm H, Eythorsson E, et al. Humoral Immune Response to SARS-CoV-2 in Iceland. New England Journal of Medicine. 2020.

10. Salzberger B, Gluck T, Ehrenstein B. Successful containment of COVID-19: the WHO-Report on the COVID-19 outbreak in China. Infection. 2020;48(2):151–3.

11. Bi Q, Wu Y, Mei S, Ye C, Zou X, Zhang Z, et al. Epidemiology and transmission of COVID-19 in 391 cases and 1286 of their close contacts in Shenzhen, China: a retrospective cohort study. Lancet Infect Dis. 2020;20(8):911–9.

12. Jing QL, Liu MJ, Zhang ZB, Fang LQ, Yuan J, Zhang AR, et al. Household secondary attack rate of COVID-19 and associated determinants in Guangzhou, China: a retrospective cohort study. Lancet Infect Dis. 2020.

13. Li W, Zhang B, Lu J, Liu S, Chang Z, Cao P, et al. The characteristics of household transmission of COVID-19. Clin Infect Dis. 2020.

14. Wang Z, Ma W, Zheng X, Wu G, Zhang R. Household transmission of SARS-CoV-2. J Infect. 2020;81(1):179–82.

15. Wu J, Huang Y, Tu C, Bi C, Chen Z, Luo L, et al. Household Transmission of SARS-CoV-2, Zhuhai, China, 2020. Clin Infect Dis. 2020.

16. Burke RM, Midgley CM, Dratch A, Fenstersheib M, Haupt T, Holshue M, et al. Active Monitoring of Persons Exposed to Patients with Confirmed COVID-19 - United States, January-February 2020. MMWR Morb Mortal Wkly Rep. 2020;69(9):245–6.

17. Rosenberg ES, Dufort EM, Blog DS, Hall EW, Hoefer D, Backenson BP, et al. COVID-19 Testing, Epidemic Features, Hospital Outcomes, and Household Prevalence, New York State—March 2020. Clinical Infectious Diseases. 2020.

18. Covid-19 National Emergency Response Center E, Case Management Team KCfDC, Prevention. Coronavirus Disease-19: Summary of 2,370 Contact Investigations of the First 30 Cases in the Republic of Korea. Osong Public Health Res Perspect. 2020;11(2):81–4.

19. Lopez Bernal J, Panagiotopoulos N, Byers C, Garcia Vilaplana T, Boddington NL, Zhang X, et al. Transmission dynamics of COVID-19 in household and community settings in the United Kingdom. medRxiv. 2020:2020.08.19.20177188.

20. Bohmer MM, Buchholz U, Corman VM, Hoch M, Katz K, Marosevic DV, et al. Investigation of a COVID-19 outbreak in Germany resulting from a single travel-associated primary case: a case series. Lancet Infect Dis. 2020;20(8):920–8.

21. Cox RJ, Brokstad KA, Krammer F, Langeland N, Bergen C-RG. Seroconversion in household members of COVID-19 outpatients. Lancet Infect Dis. 2020.

22. Silveira MF, Barros AJD, Horta BL, Pellanda LC, Victora GD, Dellagostin OA, et al. Population-based surveys of antibodies against SARS-CoV-2 in Southern Brazil. Nat Med. 2020;26(8):1196–9.

23. Di Paolo M, Iacovelli A, Olmati F, Menichini I, Oliva A, Carnevalini M, et al. False-negative RT-PCR in SARS-CoV-2 disease: experience from an Italian COVID-19 unit. ERJ Open Res. 2020;6(2).

24. Yang Y, Yang M, Shen C, Wang F, Yuan J, Li J, et al. Evaluating the accuracy of different respiratory specimens in the laboratory diagnosis and monitoring the viral shedding of 2019-nCoV infections. medRxiv. 2020:2020.02.11.20021493.

25. Zhang J, Zhang X, Liu J, Ban Y, Li N, Wu Y, et al. Serological detection of 2019-nCoV respond to the epidemic: A useful complement to nucleic acid testing. Int Immunopharmacol. 2020;88:106861.

26. Xu Y, Xiao M, Liu X, Xu S, Du T, Xu J, et al. Significance of serology testing to assist timely diagnosis of SARS-CoV-2 infections: implication from a family cluster. Emerg Microbes Infect. 2020;9(1):924–7.

27. Ludvigsson JF. Children are unlikely to be the main drivers of the COVID-19 pandemic – A systematic review. Acta Paediatrica. 2020;109(8):1525–30.

28. Stringhini S, Wisniak A, Piumatti G, Azman AS, Lauer SA, Baysson H, et al. Seroprevalence of anti-SARS-CoV-2 IgG antibodies in Geneva, Switzerland (SEROCoV-POP): a population-based study. Lancet. 2020;396(10247):313–9.

29. Grifoni A, Weiskopf D, Ramirez SI, Mateus J, Dan JM, Moderbacher CR, et al. Targets of T Cell Responses to SARS-CoV-2 Coronavirus in Humans with COVID-19 Disease and Unexposed Individuals. Cell. 2020;181(7):1489–501 e15.

